# Quality in Clinical Research: An Observational Study of Randomisation Techniques in Urological and General Surgical Studies

**DOI:** 10.1101/2023.02.15.23285955

**Authors:** Nicholas Raison, Simone Giona, Oliver Brunckhorst, Alexander Cohen, Gordon Muir

**Affiliations:** MRC Centre for Transplantation, King’s College London, London, UK; Department of Urology, Guy’s and St Thomas’ NHS Trust, London, UK; Department of Urology, Frimley Park Hospital, Camberley, UK; Department of Haematological Medicine, Guy’s and St Thomas’ Hospitals NHS Foundation Trust, London, UK; Department of Urology, King’s College Hospital, London, UK

**Keywords:** Randomised Controlled Trial, Research, Quality, CONSORT, Trials Methodology, Academic Surgery

## Abstract

**Objectives:** To quantitatively test the quality of randomised controlled trials reported to international scientific meetings through a critical analysis of randomisation outcomes.

**Design and Main Outcome Measures:** All randomised controlled trials presented at international surgical and urological congresses using simple randomisation were identified. Primary analysis of randomisation technique was performed by comparing the observed and expected numbers of trials with equal numbers of participants in each arm. Sensitivity analyses compared study design, type of study and presence of external sponsorship. All abstracts were assessed according to the CONSORT for reporting randomised trials in journal and conference abstracts checklist.

**Results:** 345 studies met the inclusion criteria. 148 studies reported simple randomisation to allocate 26,510 patients. Randomisation technique could not be identified in 104 studies. Primary and all secondary analyses demonstrated a probability of p<0.0001 that simple randomisation was used for participant allocation in all studies. Mean consort score was 9.4

**Conclusions:** It is extremely unlikely that simple randomisation was performed as reported in a significant proportion of the 148 RCTs in this study. These results raise concerning questions of the veracity and reliability of current medical research. There needs to be a greater awareness of the potential for methodological inaccuracy and error.

## INTRODUCTION

The randomised controlled trial (RCT) has long been considered the non-plus ultra of empirical research. It offers the most rigorous tool for researchers investigating the cause and effect relationship of any given collection of factors. The challenge was first set by Archie Cochrane who argued in 1972 that no new treatments should be introduced until their utility over existing treatments had been proven in a RCT(1).

Effective RCTs rely on the equal distribution of confounding factors between groups, cancelling out their effects and leaving variations in outcomes attributable only to chance or the intervention. Unbiased allocation is essential to the statistical analysis used to test for significance. RCTs are analysed on the assumption that any differences between groups will behave like differences between random samples of a single population. Importantly successful randomisation minimises the influence of investigators by ensuring that treatments or interventions cannot be predicted. The risk of substantial allocation bias, both conscious and unconscious, when investigators knowing or can predict the participants’ allocation is well recognised with outcomes exaggerated by up to 41%(2).

RCTs are not immune to fraud or deceit, particularly if not subject to overview by a third party. The estimated prevalence of fraudulent behaviour in medical research varies enormously from less than one per cent to over 25%(3). Risks from weak or erroneous trial methodologies introducing potentially profound bias into studies are thankfully now widely acknowledged. The Consolidated Statement of Reporting Trials (CONSORT) statement provides a clear minimum checklist of items that should be reported by RCTs. This has helped both investigators and readers assess the validity of studies against standard criteria. Numerous reviews have subsequently appraised various collections of studies against these standards(2, 4-7). These studies highlight the inadequate level of understanding of trial methodology which persists throughout medicine. Reporting of randomisation and allocation methods has been noted to be particularly poor(5, 6). Yet such studies remain limited by the lack of any objective assessment of methodological quality, relying instead on self-reporting by authors. The current study aims to objectively assess the quality (and potentially the veracity) of randomised controlled trials through the analysis of allocation outcomes following simple randomisation.

## Materials and Methods

The study was conducted in accordance with STROBE reporting guidelines(8). All poster presentations of surgical RCTs presented at nine international surgical conferences covering urology and general surgery between 2012 and 2017 were included in the study (full list in supplementary table 1). Two authors (NR, SG) independently identified studies before each conference through hand searches of abstract books. Studies were included if they identified themselves as randomised controlled trials in the title or abstract or reported the random allocation of patients to study arms. Studies were excluded if they did not report original data or re-analysed previously collected data; if details of randomisation or allocation were not available; if trials did not involve the randomisation of human participants or if the trial was ongoing.

A stepwise approach to data collection was undertaken. Abstracts were reviewed for details of the randomisation technique together with the numbers of patients allocated to each study arm. The CONSORT definition of simple randomisation was used. Where possible, during each conference, presentations were attended and the study authors were directly questioned as to the randomisation technique and allocation outcomes. For those cases where authors were unavailable or did not attend, they were contacted directly by email (initially the presenting author then all study authors for whom contact details were available). If no email address or contact details were provided with the poster presentation, internet searches were undertaken to find email addresses. If no email address was found or there was no response, subsequent full publications were reviewed to identify randomisation technique. All authors were then contacted to confirm the same study design was used in the abstract presented and the paper published. Studies in which randomisation details were not confirmed by any such method were excluded.

A standardised data collection template was prepared prior to data collection. Data were collected on randomisation technique; number of study arms and number of participants allocated to each arm; multicentre or single centre trial design; presence of external funding; prospective trial registration; type of trial (medical/ surgical/ mixed). “Medical trials” were defined as those assessing the use of a pharmacological agent or drug. “Surgical trials” were defined as any involving the direct assessment of a surgical intervention. Mixed trials were those that either combined a medical and surgical intervention, used a non-surgical intervention such as a catheter or involved a diagnostic intervention for example magnetic resonance imaging. All abstracts were assessed and scored according to the CONSORT for reporting randomised trials in journal and conference abstracts checklist(9). Inclusion into all public clinical trial registries was manually checked for all studies.

### Statistical Analysis

The analysis of randomisation techniques was performed by comparing the expected number of balanced studies with the observed number of balanced studies. A study was assumed to be balanced if the difference between the number of patients in each trial arm(s) was 1 or 0. Studies with unequal randomisation ratios (e.g. 2:1) were normalised to the largest group of the trial (e.g. by multiplying the smallest group with 2). The expected number of balanced studies and a one-sided p-value were calculated to test if simple randomisation was effective in each study. A total of 10,000 Monte Carlo simulations under the assumption of simple randomisation were run for each study. The expected number of balanced studies was calculated as the mean number of balanced studies per simulation cycle. One-sided p-values were calculated as the relative proportion of simulation cycles in which the number of balanced studies was equal or greater to the number of observed balanced studies. All simulations were performed using the Stata MP Version 14.2 (StataCorp, LP).

The primary analysis compared the observed and expected number of participants in all studies. Sensitivity analyses were performed by stratifying by study design, Isingle centre vs multicentre), type of study (medical/ surgical or mixed) and the presence of external sponsorship. Parametric analysis of consort scores was performed using an independent samples T-test or ANOVA as appropriate. These analyses were performed on SPSS Statistics version 25 (IBM Corp. Armonk, NY).

## Results

During the nine conferences 504 RCTs were presented of which 345 met the inclusion criteria. A specific randomisation technique was identified in 241 studies. 148 studies used simple randomisation to allocate 26,510 patients. 22 studies were classified as medical, 76 as surgical and 50 as mixed. The randomisation technique could not be identified by any method in 104 studies (Figure 1).

**Figure 1:**
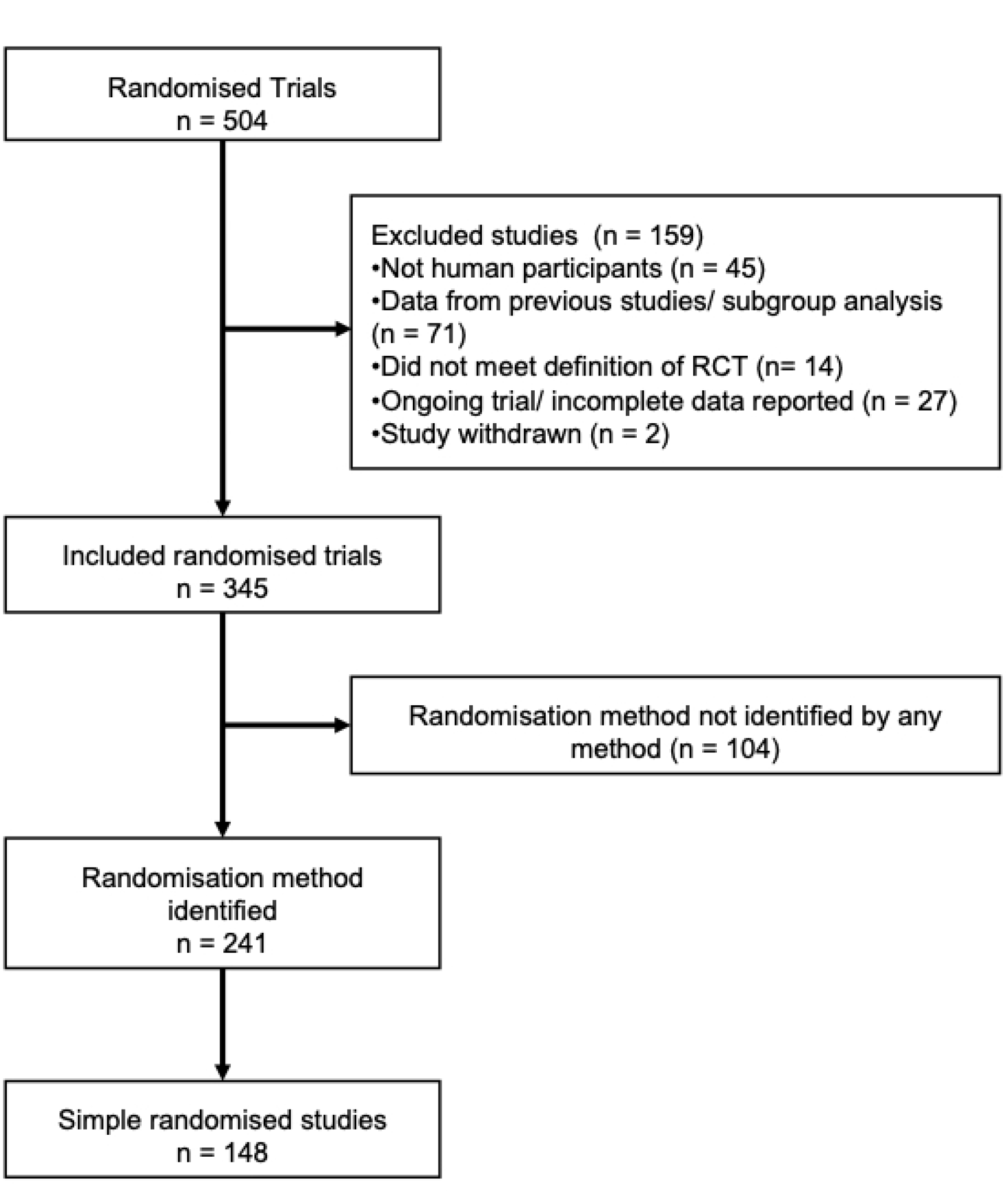
Study Flow Diagram.

Primary outcome analysis and sensitivity analyses are shown in Table 1. Results for both the primary outcome measure and all subgroup analyses demonstrate a probability of p<0.0001 that participant allocation was based only on simple randomisation in all of the included studies. Observed numbers of balanced studies greatly exceeded expected numbers in all analyses.

**Table 1:** Results of primary analysis and sensitivity analyses

| Study type |  | Expected<br>number of<br>balanced studies<br>(mean) | Observed<br>number of<br>balanced studies<br>(n) | Total<br>number of<br>studies<br>(n) | One-sided p-value |
| --- | --- | --- | --- | --- | --- |
| All studies |  | 14 | 90 | 148 | <0.0001 |
| Number of<br>centres | Multi centre | 2 | 10 | 25 | <0.0001 |
|  | Single<br>centre | 12 | 80 | 123 | <0.0001 |
| Industry | No | 10 | 74 | 119 | <0.0001 |
| sponsored? | Yes | 3 | 16 | 29 | <0.0001 |
| Type of intervention | Medical | 2 | 11 | 22 | <0.0001 |
|  | Surgical | 7 | 50 | 76 | <0.0001 |
|  | Mixed | 4 | 29 | 50 | <0.0001 |

Mean CONSORT score for all studies was 9.4. Single centre trials had significantly lower CONSORT scores than multicentre trial designs (mean CONSORT score 9.2 vs 10.3, p = 0.02). There was no difference in consort scores neither between studies with and without industry funding (mean CONSORT score 9.9 vs 9.3 p = 0.06) nor between medical, surgical and mixed studies (mean CONSORT score 9.8 vs 9.3 vs 9.2, p = 0.31). 28 studies (19%) were registered on a clinical trial registry. Registration was associated with a significantly higher CONSORT score (mean CONSORT score 10.5 vs 9.1, p < 0.0001).

## Discussion

Based on this analysis of 148 studies randomising 26,510, it is almost impossible that all studies used the simple randomisation protocol that was reported in the study methodology. No difference was seen on sensitivity analyses according to study design, presence of industrial funding or study type. Put into context, most clinical studies accept a one in twenty risk of a chance association as “significant” – here the chance is one in tens of thousands. This ignores the fact that almost one third of studies were excluded as we could not find any information on randomisation.

We also noted that a disproportionate number of studies had what we term “round numbers” of subjects (20, 50, 100, 200 etc) suggesting that no powering of the initial trial had taken place. So few reported any power calculations, and therefore may have been under powered or over powered, it was not possible to test this hypothesis.

RCTs, particularly in surgery, are complex, expensive and time consuming. Using an RCT to address a scientific or clinical questions is constrained by a number of factors. Hypotheses need to focus on relatively simple and discrete questions, frequently impractical in the clinical setting. Investigators must recruit enough participants to provide robust results, a difficult undertaken highlighted by the high numbers of underpowered studies(10). Finally, patients must be willing to undergo a randomly allocated intervention and exposure to a potentially inferior treatment must be ethically defended.

In the face of these challenges, it is therefore unsurprising that RCTs continue to suffer from poor methodologies. Systematic reviews have shown, in spite of the CONSORT guidelines, that RCTs continue to be published without critical details such as the randomisation technique(11). Reviews across various discplines have shown consistently low rates of of adequate reporting of randomisation technique: dermatology 9% (n = 6), surgery 43% (n = 65), urology 28% (n = 42) and general medicine 19% (n= 463) (5, 11-13). The quality of abstract publications have been found to be even worse prompting the release of the CONSORT statement for abstracts,. Even in high impact journals such as the Lancet, BMJ and JAMA, only 0%, 7% and 3% of abstracts respectively reported randomisation techniques(11). This study adds further evidence to support these concerning results. Despite extensive and focussed efforts by study authors, basic trial design details could not be identified in 30% of studies.

Alongside the primary outcome results demonstrating the extremely high likelihood that in many cases randomisation was performed incorrectly, numerous examples of a lack understanding were noted during interviews with presenting authors. These discussions were “off the record” and anonymous, so we cannot comment on any factors linked to those authors. A number of authors were not aware of the randomisation technique and two admitted that true randomisation was in fact not really performed! Other studies reported techniques such as patient preference, clinician availability and simply alternating between techniques. Quasi-randomisation methods such as according to the date of birth, hospital record number or day of the week were used in a number of studies. In all cases the studies reported using simple randomisation.

Interpretation of these results must be considered in the context of a number of limitations. We relied on the presenting authors to understand and accurately report the randomisation technique. In some cases, junior authors presenting the studies were clearly unfamiliar with the details of the trial methodology and may have described the randomisation technique incorrectly. Including only poster presentations rather than full publications in peer reviewed journals may also have led to the inclusion of a greater proportion of low-quality studies. However previous attempts by the current authors to gather randomisation data from authors of published studies garnered response rates of less than 10%. As result the direct approach of questioning authors at their posters was felt to be justified. These results focus on urological studies as it is the specialty of the majority of the authors. As in previous work on CONSORT compliance, the study was extended to include general surgical and non-urological specialties(6). Although direct comparisons were not performed, results with high proportions of balanced studies were seen in both urological and non-urological studies. Likewise, author interviews revealed comparable numbers of suspect answers. Replication of this research in other disciplines would be welcome. Finally given the nature of the analysis undertaken in this study, it is only possible to report the overall likelihood that simple randomisation was performed as reported. No assessment can be made on the exact numbers of studies that reported erroneous methodologies.

This study included only RCTs that used simple randomisation. Simple randomisation represents the most straightforward approach to allocating patients but can still be difficult to implement (14). All participants should have an equal chance of being allocated to either arm. Especially with smaller sample sizes, this can result in unequal group sizes and important factors may be not be balanced between groups. Yet the false belief that samples sizes should be equal remains pervasive(7, 15). This misconception is reflected in our results with 59% of studies (n=88/148) reporting equal arms or differing by just 1.

Despite these drawbacks, RCT’s remain a cornerstone for the advancement of medical practice. The COVID-19 pandemic has clearly shown the importance of high quality RCTs with their results directing clinical management and policy across the world.

However often in everyday clinical practice, especially in surgery, such sophisticated study designs are potentially being overused. The value of well conducted, non-randomised studies is often overlooked by both researchers and funding bodies. This study has shown that in many cases, RCT’s are beyond the abilities of surgical teams. Sadly medical history is littered with poorly executed and fraudulent studies whose biased results have misled medical opinion, occasionally for decades. Alarming figures for the prevalence of research misconduct and rising numbers of retractions have prompted many national government to appoint national bodies to regulate and provide oversight on academic research(16). The results from this study suggest similar practises continue in academic research. Whilst a proportion of errors will be attributable to a lack of experience and ability of the study authors, wilful dishonesty must also be considered as factor. The common conclusion to so many non-randomised studies that an RCT is required to provide the definitive answer must be reviewed. Instead the utility of well-designed comparative studies should be better advocated. For the majority of smaller studies, a non-randomised trial design would help greatly simplify the studies, enabling surgical groups focus on achieving true and reliable results.

## Conclusion

This study has shown that it is extremely likely that for a significant proportion of 148 simple randomised studies, randomisation was not performed as described. All studies had been selected for presentation following a peer review process and their results reported in international scientific journals. These results raise important questions on the current quality of medical research and reliability of clinicians to undertake research. We cannot say why these results have occurred – ignorance of the RCT process may be the reason, yet we know from personal experience that the majority of institutions presenting data have taken part in high quality multicentre RCT’s which would not fall foul of our scrutiny. Pressure to publish may mean that, particularly for single centres studies, corners were cut and proper protocols were ignored for simplicity. All trials must be registered prospectively with clear, publicly disseminated randomisation protocols. Of course, fraud is an ever-present explanation for research findings which are either too good to be true or which do not make sense. Faced with the above however, it would seem that any single centre RCT submitted is likely to be greatly at risk of basic statistical flaws – we would suggest that editors would be well advised to seek a statistician’s advice before sending such trials out to hard pressed reviewers.

## Data Availability

All relevant data are within the manuscript and its Supporting Information files

## Ethical approval

Ethical approval not required.

## Contributorship Statement

All authors meet the criteria for authorship

## Guarantor

The guarantor, Mr G Muir, takes responsibility for the integrity of the work and confirms that he controlled the decision to publish.

## Acknowledgements

We thank Stephen Rietbrock and Christopher of the Wallenhorst, Institute for Epidemiology, Statistics and Informatics for performing the statistical analysis. Neither were received compensation for their role in the study

## Supplementary Table Legends

Supplementary Table 1: Surgical and Urological Conferences Included in this Study

## References

1. Cochrane AL. Effectiveness and efficiency: random reflections on health services. London: Nuffield Provincial Hospitals Trust; 1972. xi, 92 p. p.

2. Schulz KF, Chalmers I, Hayes RJ, Altman DG. Empirical evidence of bias. Dimensions of methodological quality associated with estimates of treatment effects in controlled trials. JAMA. 1995;273(5):408–12.

3. Sarwar U, Nicolaou M. Fraud and deceit in medical research. J Res Med Sci. 2012;17(11):1077–81.

4. Turner L, Shamseer L, Altman DG, Weeks L, Peters J, Kober T, et al. Consolidated standards of reporting trials (CONSORT) and the completeness of reporting of randomised controlled trials (RCTs) published in medical journals. Cochrane Database Syst Rev. 2012;11:MR000030.

5. Adie S, Harris IA, Naylor JM, Mittal R. CONSORT compliance in surgical randomized trials: are we there yet? A systematic review. Ann Surg. 2013;258(6):872–8.

6. Agha R, Cooper D, Muir G. The reporting quality of randomised controlled trials in surgery: a systematic review. Int J Surg. 2007;5(6):413–22.

7. Schulz KF, Chalmers I, Grimes DA, Altman DG. Assessing the quality of randomization from reports of controlled trials published in obstetrics and gynecology journals. JAMA. 1994;272(2):125–8.

8. von Elm E, Altman DG, Egger M, Pocock SJ, Gotzsche PC, Vandenbroucke JP, et al. The Strengthening the Reporting of Observational Studies in Epidemiology (STROBE) statement: guidelines for reporting observational studies. J Clin Epidemiol. 2008;61(4):344–9.

9. Hopewell S, Clarke M, Moher D, Wager E, Middleton P, Altman DG, et al. CONSORT for reporting randomised trials in journal and conference abstracts. Lancet. 2008;371(9609):281–3.

10. Dimick JB, Diener-West M, Lipsett PA. Negative results of randomized clinical trials published in the surgical literature: equivalency or error? Arch Surg. 2001;136(7):796–800.

11. Hays M, Andrews M, Wilson R, Callender D, O’Malley PG, Douglas K. Reporting quality of randomised controlled trial abstracts among high-impact general medical journals: a review and analysis. BMJ Open. 2016;6(7):e011082.

12. Scales CD, Jr., Norris RD, Keitz SA, Peterson BL, Preminger GM, Vieweg J, et al. A critical assessment of the quality of reporting of randomized, controlled trials in the urology literature. J Urol. 2007;177(3):1090-4; discussion 4-5.

13. Adetugbo K, Williams H. How well are randomized controlled trials reported in the dermatology literature? Arch Dermatol. 2000;136(3):381–5.

14. Kao LS, Tyson JE, Blakely ML, Lally KP. Clinical research methodology I: introduction to randomized trials. J Am Coll Surg. 2008;206(2):361–9.

15. Altman DG, Dore CJ. Randomisation and baseline comparisons in clinical trials. Lancet. 1990;335(8682):149–53.

16. Tavare A. Managing research misconduct: is anyone getting it right? BMJ. 2011;343:d8212.

